# Implementation of a Statewide Endovascular Aortic Aneurysm Surveillance Program Improves One-Year Follow-up and Survival

**DOI:** 10.1101/2025.06.20.25330024

**Authors:** Frank M. Davis, Jeremy Albright, Brennan Callow, Loay Kobbani, Hue Thai, Tamer Boules, Nicolas J. Mouawad, Nicholas Osborne

## Abstract

**BACKGROUND:** National guidelines recommend long-term surveillance after endovascular AAA repair (EVAR) to monitor for endoleaks, graft migration or device failure. However, investigations have demonstrated that adherence to EVAR surveillance can vary greatly. Recently, the BMC2 Cardiovascular Consortium (BMC2) implemented a statewide performance metric, linked with financial reimbursement, designed to improved EVAR postoperative surveillance. The purpose of this study was to evaluate the impact of this statewide metric on EVAR follow-up and long-term endovascular outcomes.

**METHODS:** The BMC2 statewide cardiovascular consortium consisting of 36 hospitals was queried for patients who underwent EVAR between 2017-2023. Patient, hospital and operative risk factors were collected and analyzed based on the presence or absence of follow-up EVAR surveillance imaging. The primary outcome was adherence to recommended EVAR surveillance and the longitudinal change in percent of EVARs that obtained surveillance imaging.

**RESULTS:** A total of 5,742 patients underwent EVAR between 2017-2023. During the implementation of the EVAR surveillance metric, there was steady increase in the annual percentage of EVARs across the state who received postoperative 1-year EVAR surveillance imaging (26.46% in 2017 to 77.69% in 2023). On mixed logistic modeling, predictors of incomplete post-EVAR 1-year surveillance included emergent operation, length of stay and non-home discharge. For those patients that received appropriate EVAR surveillance imaging follow-up they displayed a higher number of repeat interventions (2.7% vs.1.6%; p<0.005) but had a decrease in one-year mortality (0.7% vs 10.6%; p<0.001) compared to those did not receive 1-year EVAR surveillance.

**CONCLUSIONS:** Incomplete imaging surveillance following EVAR remains high and predisposes patients to worse survival outcomes. Utilization of a performance metric both with associated financial incentive, within a statewide quality collaborative is a viable strategy that can have a significant impact on increasing EVAR surveillance and decreasing one-year mortality after endovascular AAA repair.

## INTRODUCTION

Endovascular aneurysm repair (EVAR) has progressively replaced open aneurysm repair over the past two decades for the treatment of infrarenal abdominal aortic aneurysms (AAAs).^1^ Although peri-operative morbidity and mortality following EVAR are low, significant concerns exist regarding the long-term durability of EVAR with high rates of secondary interventions, with a cumulative incidence of 14% at 5 years.^2^ Further, endoleak is the most frequent complication after EVAR, seen in up to 25% of patients during follow-up which can progress to late aortic rupture with an incidence of 1.5%.^1,3^ As such EVAR requires active long-term surveillance for the detection and correction of late endograft complications. Current, Society for Vascular Surgery practice guidelines recommend lifelong surveillance with computed tomographic (CT) scanning or color duplex at 1 and 12 months during the first postoperative year, followed by repeat imaging every 12 months thereafter, so that timely re-intervention can prevent late aneurysm rupture.^4^

Compliance with post-EVAR imaging surveillance remains suboptimal. A recent large comparative meta-analysis demonstrated that on average 42% of EVAR patients fail to meet current guidelines.^5^ Additionally, inadequate long-term EVAR surveillance has been shown to be as high as 57% in Medicare beneficiaries^6^ and 51% of Veterans Affairs beneficiaries.^7^ Although some reports have called the utility of annual post-EVAR surveillance into question^6,8^, multiple investigation have demonstrated that inadequate long-term surveillance after endovascular repair results in an increase in all-cause patient mortality and aneurysm related mortality at 5 years.^9–13^ As a consequence, different strategies, both in person and remote, have been employed in an attempt to increase EVAR follow-up but none have demonstrated marked improvement in surveillance imaging or long-term outcomes.^9^

Within this context, in 2017 a cardiovascular quality improvement collaborative within the state of Michigan (BMC2 Cardiovascular Consortium) implemented a statewide reporting mechanism for EVAR surveillance. BMC2 as a quality improvement platform recognizes hospital performance using a Pay-for-Performance (P4P) model. This program is administered by Blue Cross Blue Shield of Michigan to pay hospitals in aggregate an additional 5% of inpatient and outpatient operating payments. The goals for the inputs (quality metrics) to this program are determined in concert with the quality collaboratives (i.e. BMC2) two years in advance (https://www.bcbsm.com/amslibs/content/dam/public/providers/documents/value/pay-for-performance-program-guide-2024-2025.pdf). As part of the statewide quality improvement program, BMC2 also implemented a P4P goal to increase the rate of post-EVAR 1-year surveillance imaging beginning in 2021 (reported in 2022) with a goal of 70% of patients within each hospital obtaining imaging between 6 months and 14 months following EVAR. This goal was announced in 2020 in anticipation of the P4P. The goal has since been increased to 80% compliance in 2024 (https://bmc2.org/quality-improvement/performance-indexes). Within this study, we investigated the impact of this statewide cardiovascular quality improvement program on altering the rate of long-term EVAR follow-up. We specifically evaluated annual EVAR surveillance adherence rates and the predictors for noncompliance before and after program initiation as well as the impact of EVAR surveillance on secondary interventions and patient mortality.

## METHODS

### Data Source

The BMC2 Cardiovascular Consortium (BMC2) Vascular Surgery Intervention Resgistry ), a prospective, multicenter observational registry and quality improvement program, served as the data source for this study. The details of the BMC2 program have been previously described.^14–16^ Trained nurses or clinical abstractors entered data prospectively on >100 clinical and demographic variables. Research analysts were blinded to patient, surgeon, and hospital identity. As part of the BMC2, quality improvement projects have focused on actionable opportunities to improve patient outcomes. Some of these quality improvement initiatives have been included in a Pay-for-Performance (P4P) plan to promote improvement across hospitals in the state. Based upon hospital performance across multiple metrics, hospitals are entitled to a financial incentive for meeting performance goals. Within this study, in 2017, BMC2 implemented the quality improvement performance goal post-EVAR 1-year surveillance imaging in at least 70% of patients who underwent an endovascular repair. Between 2017-2023 we monitored the rate of post-EVAR 1-year surveillance imaging. The University of Michigan School of Medicine IRB has determined that study using BMC2 is exempt under 45 CFR 46.101(b)(4) and thereby a waiver for informed consent was granted. *Support for BMC2 is provided by Blue Cross Blue Shield of Michigan (BCBSM) and Blue Care Network as part of the BCBSM Value Partnerships program. Although BCBSM and BMC2 work collaboratively, the opinions, beliefs, and viewpoints expressed by the authors do not necessarily reflect the opinions, beliefs, and viewpoints of BCBSM or any of its employees. Further, BCBSM does not have access to BMC2 data, and all patient episodes occurring at engaged hospitals are included in the data registries, regardless of payer*.

### Cohort of Patients

The study analyzed all patients undergoing an elective infrarenal EVAR of an AAA between January 2017 and January 2023 (follow-up included through 2024). We excluded physician modified graft configurations and patients who received snorkels or chimney stents as a means for visceral revascularization. Patients who underwent repair for a thoracic aortic aneurysm, thoracoabdominal aneurysms, or aortic dissection were also excluded from the study. Over this period, 5,742 elective EVARs were conducted. There are 36 healthcare systems included in the BMC2 however, only 20 healthcare systems performed at least one elective EVAR per year during the study period and as such were included for analysis in this study. It should be noted that all healthcare systems analyzed in this study also participated in the VQI registry.

Baseline characteristics included patients’ demographics, comorbid conditions, and anatomic and procedural characteristics. Regarding the BMC2 Vascular Surgery data collection, data abstractors collect and enter all data for each patient, including demographic information, past medical history, standard laboratory test results, technical details of procedures, early and late outcomes, including associated complications, if they occurred. Data quality is controlled at 3 levels starting with manual review at the point of data entry to insure completeness and validity by trained data abstractors. A database manager confirms the quality of data entry and analyzes random samples of data for clinical consistency. Finally, twice per year a nurse coordinator from BMC2 performs a site visit to audit all cases with severe complications and a random sample of 5% of cases.

### Outcomes

The primary outcome in this study was the rate of adherence to Society of Vascular Surgery EVAR practice guidelines with documented post-EVAR 1-year surveillance imaging. Given the requirements for P4P, BMC2 has defined post-EVAR 1-year surveillance imaging as either CT or color duplex imaging performed between 6 months and 14 months. Secondary outcomes were analyzed including the need for secondary intervention and patient mortality.

Re-intervention was defined on an intention to treat basis, and all participating centers followed a policy of endograft re-intervention in cases of demonstrable type 1 or 3 endoleak, sac expansion or device migration >5 mm on cross sectional imaging, or in the presence of symptomatic endograft limb stenosis or occlusion. Type 2 endoleak was subject to re-intervention only if associated with sac expansion. Data quality and the inclusion of consecutive procedures are ensured by ad hoc queries, random chart review, and a series of diagnostic routines included in the database conducted by the coordinating center. Coronary artery disease was defined as a history of myocardial infarction, percutaneous coronary intervention, or coronary artery bypass graft. Cerebrovascular disease was defined as a history of stroke or transient ischemic attack.

### Statistical Analysis

Patients were categorized into those with and without imaging surveillance performed at 1 year follow-up and compared on demographic, clinical history and procedural variables. Categorical variables are presented as count and percentage and continuous variables as with means and standard deviations or, if skewed, medians and interquartile ranges. Differences in characteristics between groups were compared using χ^2^ or Fisher exact tests where appropriate for categorical variables, and t-tests or Wilcoxon-Mann-Whitney tests were used for continuous variables. No adjustments in data analysis were made during the period of the COVID pandemic.

Multivariate analysis was performed using Bayesian mixed effects logistic regression. Each model included a random effect for hospital along with default diffuse priors for all parameters. Odds ratio estimates with 95% highest posterior credible intervals bounded away from 1.00 are considered significant. All calculations were performed with the statistical software R (version 3.0.2), and the rstanarm package (version 2.21.4) was used for the mixed models.

## RESULTS

The study population consisted of 5,742 patients (19.3% women) who underwent elective endovascular AAA repair (EVAR) at centers across the state of Michigan between 2017-2023. The average age was 73 (standard deviation, 8.9) years. Approximately, 92% of the study population were white, and 8% were black or of another race. Vascular-related comorbidities were highly prevalent across the study population, including hypertension (87%), hyperlipidemia (87%), coronary artery disease (47%), and chronic obstructive pulmonary disease (43%). The mean BMI was 28.4 (SD = 6.0), and 37% were current smokers (Table I). Of the 5,742 patients who underwent elective EVAR repair during the study period, 2,801 patents (48.8%) failed to have post-EVAR 1-year surveillance imaging by CTA or ultrasound imaging. Unadjusted bivariate analysis showed that compared to the patients who obtained 1-year surveillance imaging, patients without imaging follow-up had a higher rate of CAD (49.3% vs. 45.3%, p=0.003), congestive heart failure (17.8 vs. 15.2%; p=0.008), and anemia (28.0% vs. 23.0%, p=0.001). Further when analyzing the index hospitalization, those patients that did not have follow-up surveillance imaging had a higher rate of postoperative complications during the index procedure including respiratory failure (0.8% vs. 0.2%; p=0.006). Patients with follow-up surveillance were more likely to have a non-home discharge, including to a nursing facility, after an EVAR (6.8% vs. 3.2%, p < 0.001) (Table I). We then conducted an adjusted analysis to determine predictors of incomplete post-EVAR 1-year surveillance imaging. On mixed logistic modeling, post-procedure length of stay and patient non-home discharge were significantly associated with lack of post-EVAR 1-year surveillance imaging. In contrast, hyperlipidemia was found to be associated with appropriate 1-year EVAR surveillance (Table II).

**Table I.** Baseline demographics and clinical history of patients.

| Variable | Overall<br>(n=5,742) | Surveillance<br>Imaging Performed<br>(n=2,941) | No Imaging<br>Performed<br>(n=2,801) | *P Value* |
| --- | --- | --- | --- | --- |
| ***Demographics*** |  |  |  |  |
| Age | 72.70 (8.85) | 72.57 (8.75) | 72.84 (8.95) | 0.243 |
| Sex = Female (%) | 1,110 (19.3) | 549 (18.7) | 561 (20.0) | 0.203 |
| Race (%) |  |  |  | 0.071 |
| White | 5,291 (92.1) | 2,731 (92.9) | 2,560 (91.4) |  |
| Black | 284 (4.9) | 127 (4.3) | 157 (5.6) |  |
| Other | 167 (2.9) | 83 (2.8) | 84 (3.0) |  |
| BMI | 28.38 (6.04) | 28.56 (6.15) | 28.20 (5.92) | 0.023 |
| Ever Smoked (%) | 5,197 (90.5) | 2,652 (90.2) | 2,545 (90.9) | 0.399 |
| Current Smoker (%) | 2,135 (37.2) | 1,078 (36.7) | 1,057 (37.7) | 0.412 |
| Hyperlipidemia (%) | 4,974 (86.6) | 2,579 (87.7) | 2,395 (85.5) | 0.017 |
| Hypertension (%) | 4,994 (87.0) | 2,537 (86.3) | 2,457 (87.7) | 0.110 |
| Diabetes Mellitus (%) | 1,282 (22.3) | 642 (21.8) | 640 (22.8) | 0.370 |
| CAD (%) | 2,711 (47.2) | 1,331 (45.3) | 1,380 (49.3) | 0.003 |
| COPD (%) | 2,484 (43.3) | 1,224 (41.6) | 1,260 (45.0) | 0.011 |
| Renal Failure (%) | 55 (1.0) | 24 (0.8) | 31 (1.1) | 0.320 |
| Anemia (%) | 1,460 (25.4) | 677 (23.0) | 783 (28.0) | 0.001 |
| Significant Valve Disease (%) | 505 (8.8) | 240 (8.2) | 265 (9.5) | 0.091 |
| CHF (%) | 944 (16.4) | 446 (15.2) | 498 (17.8) | 0.008 |
| Prior Aortic Surgery (%) | 274 (4.8) | 130 (4.4) | 144 (5.1) | 0.223 |
| Top Deprivation Quintile (%) | 769 (13.6) | 349 (12.0) | 420 (15.2) | <0.001 |
| ***Procedure Characteristics*** |  |  |  |  |
| Max AAA Diameter (mm) | 57.55 (12.27) | 57.76 (12.87) | 57.35 (11.67) | 0.217 |
| Procedure Length (Hrs) | 4.09 (116.07) | 5.62 (165.55) | 2.64 (14.28) | 0.331 |
| Estimated Blood Loss (mL) | 75.00 [50.00, 150.00] | 100.00 [50.00, 150.00] | 69.00 [40.00, 150.00] | 0.002 |
| Respiratory Failure (%) | 29 (0.5) | 22 (0.8) | 7 (0.2) | 0.006 |
| Post-op MI (%) | 22 (0.4) | 14 (0.5) | 8 (0.3) | 0.237 |
| Post-op Infection/Sepsis (%) | 38 (0.7) | 28 (1.0) | 10 (0.3) | 0.004 |
| Post-op Renal Failure (%) | 15 (0.3) | 13 (0.5) | 2 (0.1) | 0.007 |
| Length of Stay (days) | 1.00 [1.00, 2.00] | 1.00 [1.00, 2.00] | 1.00 [1.00, 1.00] | <0.001 |
| Non-Home Discharge (%) | 285 (5.0) | 191 (6.8) | 94 (3.2) | <0.001 |

BMI, Body mass index; CAD, Coronary artery disease; CHF, congestive heart failure; COPD,
Chronic obstructive pulmonary disease; MI, myocardial infarction
Continuous variable are summarized by mean (standard deviation) and categorical variables are summarized by N (%). P values are calculated from the t-test for continuous variables and $\chi^2$ test for categorical variables.

**Table II.**
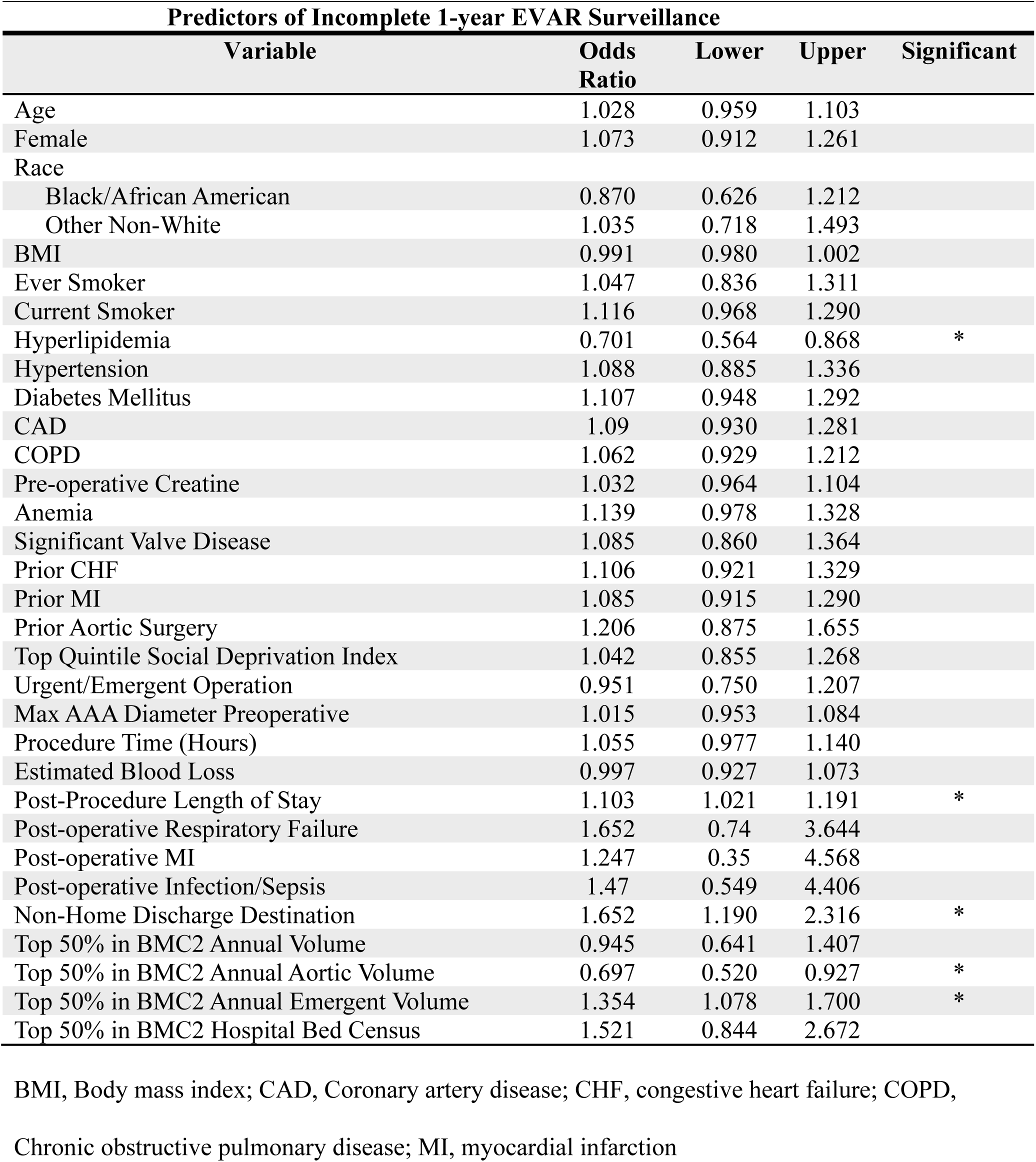
Predictors of Incomplete 1-year EVAR Surveillance Predictors of Incomplete 1-year EVAR Surveillance.

### Statewide hospital variation in 1-year EVAR surveillance

Analysis of EVAR surveillance by year demonstrated that prior to implementation of the statewide program, only 26.46% of patients who had an EVAR in 2017 had documented post-EVAR 1-year surveillance imaging and the rate of EVAR surveillance varied greatly by hospital from 3.45% of patients at the lowest hospital to 62.5% of EVAR patients at the highest hospital (Figure 1A, B). To gain insight into the variation of EVAR imaging surveillance, we analyzed hospital characteristics for each of the medical centers participating in the vascular collaborative. Hospitals were divided into low and high imaging surveillance rates based on the percentage of patients who had an EVAR that received post-EVAR 1-year surveillance imaging. Interestingly, hospitals with a higher percentage of emergent operations were more likely to have inadequate 1-year surveillance imaging follow-up while those with an annual aortic volume within the top 50% of the state were more likely to have appropriate 1-year imaging follow-up (Table II).

**Figure 1.**
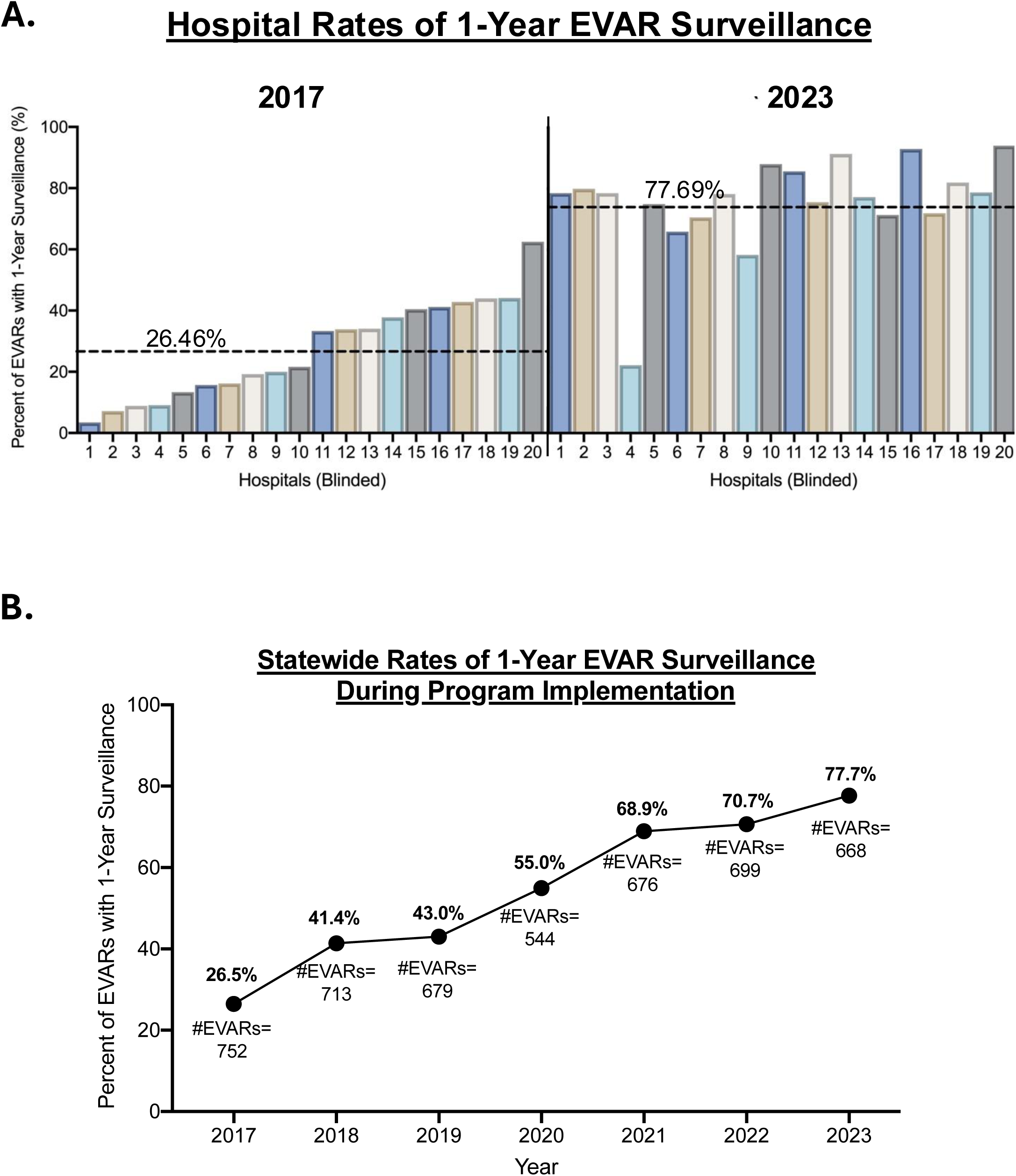
Rate of 1-year EVAR Surveillance across the statewide BMC2. **A.** Percentage of patients who receive 1-year EVAR surveillance by hospital system in 2017 and 2023. **B.** Change in the percentage of patients who receive 1-year EVAR surveillance across the entire state from 2017-2023.

### Impact of Statewide EVAR Surveillance Program

To augment the rate of EVAR surveillance, the BMC2 Cardiovascular Consortium Vascular Surgery (BMC2) implemented a vascular surgery performance metric, linked to post-EVAR 1-year surveillance imaging. Specifically, one of the pay-for-performance metrics required that hospitals document post-EVAR 1-year surveillance imaging in at least 70% of their EVARs. Following initiation of this program in 2017, there was a steady increase in the percentage of patients who underwent EVAR surveillance at 1 year (Figure 1A, B). By 2023, 77.69% of patients, averaged across all participating hospitals, who underwent an EVAR had surveillance imaging performed at 1-year timepoint after their index operation. Further by 2023, eighteen of the twenty hospitals across the collaborative achieved rates of EVAR follow-up >60%. This is a substantial improvement in comparison to 2017 where there was only one hospital within the collaborative that had an EVAR follow-up rate of >60% (Figure 1A).

### Impact of EVAR surveillance on morbidity and mortality

To analyze the impact of the statewide EVAR surveillance program on patient morbidity and mortality, a mixed logistic model was constructed to first analyze reintervention. Consistent with previous investigations, maximum AAA diameter at the time of operative intervention was a significant predictor of need for reintervention following EVAR (OR 1.546; 95% interval: 1.252-1.913). In addition, adherence to post-EVAR 1-year surveillance imaging was also found to be a predictor for need for reintervention (OR 1.829; 95% interval: 1.187-2.834; Table III and Supplemental Figure 1). Thereby suggesting that EVAR surveillance was able to adequately detect EVAR durability concerns requiring secondary intervention. We further performed a multivariate analysis to determine independent patient, procedural, or hospital factors associated with one-year mortality following EVAR and found that high risk medical commodities, including pre-operative creatine, age, significant valvular disease, anemia and congestive heart failure, were all independently associated with increased risk of 1-year mortality following EVAR. However, the factor that was found to be associated with the lowest 1-year mortality was if post-EVAR 1-year surveillance imaging was performed (OR 0.051; 95% interval: 0.029-0.085; Table III and Supplemental Figure 2).

**Table III.** Predictors of Reintervention and One Year Mortality.

| **Reintervention** |  |  |  |
| --- | --- | --- | --- |
| **Variable** | **Odds Ratio** | **Lower** | **Upper** |
| Black/African American | 2.620 | 1.033 | 5.992 |
| Surveillance Imaging Performed | 1.829 | 1.187 | 2.834 |
| Maximum AAA Diameter | 1.546 | 1.252 | 1.913 |
| Top Deprivation Quintile (%) | 0.375 | 0.155 | 0.796 |

| **Mortality** |  |  |  |
| --- | --- | --- | --- |
| **Variable** | **Odds Ratio** | **Lower** | **Upper** |
| Surveillance Imaging Performed | 0.051 | 0.029 | 0.085 |
| BMI | 0.967 | 0.942 | 0.992 |
| Pre-Op Creatinine | 1.135 | 1.011 | 1.269 |
| Length of Stay | 1.257 | 1.090 | 1.446 |
| Age | 1.531 | 1.295 | 1.816 |
| Significant Valve Disease | 1.578 | 1.040 | 2.353 |
| Anemia | 1.727 | 1.281 | 2.327 |
| CHF | 1.854 | 1.313 | 2.607 |

## DISCUSSION

Long-term durability has become one of the most important differences between open aneurysm repair and EVAR. Contemporary research suggests that, unlike open abdominal aortic aneurysm repair, EVAR requires vigilant post implantation monitoring to rescue late device failure.^17–19^ As such, current cardiovascular guidelines advise surveillance with computed tomography scans or contrast ultrasound at 1 month and 12 months postoperatively and then annually thereafter to detect endoleaks and aneurysmal enlargement.^4^ However, prior investigations demonstrated a large variability to adequate follow-up and called the utility of these guidelines into question. In the current study, we demonstrate that implementation of a statewide quality collaborative and associated performance metric can profoundly increase the rate of EVAR postoperative surveillance at both the individual patient level and within healthcare systems. We further show that those patients who received surveillance imaging displayed increased number of reinterventions but decreased one year mortality independent of patient risk factors.

Within this study, during the initial years of the program implementation, compliance with guideline EVAR surveillance was low and varied widely across health care systems in the state. Indeed in 2017 on average only 26.46% of patients who had an EVAR went on to have post-EVAR 1-year surveillance imaging. Further when analyzing by healthcare system the rates of EVAR surveillance varied from 3.57%-62.50%. This low rate of EVAR surveillance has been previously shown in a metanalysis by Grima et al. which incorporated thirty-one studies analyzing EVAR follow-up and concluded that compliance rates with EVAR guidelines was only 38% (95% CI 27-48%).^5^ Additionally, the variability in compliance across healthcare system has also been documented as a multicenter analysis from the United Kingdom with 1,414 elective EVAR patients across 10 centers demonstrated that compliance varied from 9% to 88% between centers.^13^ In an effort to improve the rate of post-EVAR surveillance hospitals have participated in quality improvement registries (such as the Vascular Quality Improvement registry) but this has not demonstrated a sustainable improvement in post-EVAR surveillance metrics.^20^ As such new strategies are urgently warranted to improve post-EVAR surveillance imaging. Unlike other studies, our study implemented a program specifically designed to improve the rate of one-year EVAR surveillance and adherence to AAA treatment guidelines across the region. Within an existing statewide quality improvement program, we have shown that both measuring hospital EVAR surveillance rates and tethering these rates to a pay-for-performance goal can result in substantial improvement in guideline-appropriate EVAR follow-up. With EVAR surveillance rates increasing from 26.46% in 2017 to 77.69% in 2023, this is the first study that has been designed and implemented a program that can markedly increase guideline appropriate imaging follow-up. Importantly, this average change in EVAR follow-up was not driven by a few selective high performing healthcare systems. Instead, we observed uniform increase in rates of EVAR surveillance regardless of hospital involved such that by 2023 eighteen of hospitals with the cohort (80%) had EVAR surveillance above 60% which is in comparison to 2017 at which time one hospital (5%) within the cohort had a follow-up rate above 60%.

Over the years of its existence, the BMC2 Cardiovascular Consortium has implemented multiple quality improvement projects linked with a Pay-for-Performance plan to promote implementation of best practice guidelines across hospitals in the state. Importantly, improvement began prior to the implementation of the P4P goal during the initial reporting phase (2017-2021). Subsequent improvement in 1 year imaging surveillance has been consistent across nearly all hospitals, showing the power of both reporting outcomes, but also incentivizing improvement. Prior investigations on the use of financial incentives to improve delivery of health care have been mixed.^21–23^ As recent reviews of pay-for-performance programs found evidence of short-term improvements in some care processes, such as guideline-recommended screening and prescribing, but no impact on longer-term patient outcomes.^21,23^ Additionally, systematic reviews observed that incentive programs appear more likely to show desired effects for process rather than outcome measures, if desired behaviors are specific and easy to measure and are in areas where there is clear room for improvement.^22^ This is in line with social psychology experiments in fields outside medicine that have established that financial carrots work as expected for mechanical, repetitive activities where higher rewards generally stimulate greater effort and more of the desired output. However, for more complex activities that require greater cognitive input, financial incentives often lead to poorer performance. This counterintuitive result is attributable to several unintended effects that incentives can have on behavior including limiting intrinsic motivation and lead to uncooperative behavior. Perhaps most damaging of all, incentives can be highly addictive, leading to net deteriorations in quality following their withdrawal.^24^ In order to maintain the long-term success of the EVAR surveillance program we are currently working to identify the processes of care at each participating hospital that allowed the surveillance program to become “mechanical and repetitive” for the healthcare teams such as use of virtual follow-up, automated physician and nursing reminders interrogated into the electronic medical record, or automated patient calls to remind them for their imaging appointments. In doing so we will work to identify the forces that drove the success of this program so that it can be implemented more broadly.

When analyzing the impact of improved EVAR surveillance our study specially investigated rates of reintervention and mortality. We found that long-term imaging follow-up was associated with increased reinterventions. This finding is consistent with prior single center studies, large Medicare populations investigations, and one metanalysis which have all reported long term EVAR surveillance increases the rate of major and minor reintervention. ^6,12,25^ Separately we examined whether there is an association between patient surveillance imaging and postoperative survival. Currently the literature is divided on the impact of EVAR imaging surveillance on long-term patient mortality. In a review of 9503 Medicare patients, Garg et al. demonstrated that nonadherence to EVAR surveillance imaging guidelines was not associated with worse aneurysm related mortality or long-term mortality at 5 years. Instead, patients with incomplete follow-up demonstrated a decrease in all-cause mortality.^6^ Similar findings with decreased mortality in a nonadherence group have been shown using smaller institutional cohorts while other large cohort studies have demonstrate no difference in mortality between patients with adequate and inadequate surveillance.^10,25,26^ The authors hypothesize that this paradoxical finding in which surveillance increases the risk of all-cause mortality may be due to patients with other considerable medical conditions that increase the risk for death may be more likely to have physician visits and undergo surveillance. In contrast to these studies, a large study of 11,309 elective EVARs within the Vascular Quality Initiative data set demonstrated that routine in-person follow-up and imaging after EVAR was associated with significant decrease in long-term mortality.^9^ This finding has also been replicated in a smaller single center study.^12^ This is in agreement with our investigation in which the data suggest that patients with inadequate follow-up imaging have inferior long-term outcomes and increased mortality compared to patients who have documented imaging surveillance. Although our study noted differences in baseline medical comorbidities between the groups, after controlling for baseline differences the risk of one year mortality was significantly less in patients who received appropriate EVAR surveillance. Further investigations are necessary to see if the impact of this statewide collaborative EVAR surveillance metric persists on 5- and 10-year patient survival. This thereby supports the importance of EVAR follow-up to monitor EVAR durability and prevent potential complications.

Our study is subject to limitations of large database studies. Compared to administrative data sources, the BMC2 data is unlikely to have significant coding errors since all patient records are manually abstracted by trained and audited abstractors.^27^ Importantly, we cannot account for all potential confounders (such as the travel distance from the patient’s home to treating hospital or if the patient had follow-up with a surgeon other than the operating surgeons) despite rigorous standardized prospective collection and assessment of data and statistical adjustments. Follow-up can also be underestimated if patients obtain imaging follow-up outside of the captured Electronic Health Record. Abstractors are trained to use linking records such as Epic CareEverywhere^TM^, but it is possible that some patients are obtaining in systems that do not participate. Efforts within BMC2 have focused on expanding the capture of all patient records. Finally, our dataset derives from patients within the State of Michigan where we have a dominant insurance provider and a networked group of hospitals participating in a quality collaborative with financial incentives and as a result may not be generalizable outside of the state of Michigan because of the differences in hospital characteristics, insurance payers, surgical practice, or access to relevant resources. However, lessons learned from this unique situation may be able to be utilized in other arenas to improve the rate of EVAR surveillance. For many centers within the statewide collaborative have harnessed virtual or remote follow-up appointments to allow for patients to obtain CT scans at institutions closer to their home while also coordinating with the initial treatment hospital/physician to confirm absence of endoleaks within the EVAR. Additionally, the mix of academic, private, and community hospitals within this study represents the pragmatic, real world experience. We do not believe these limitations detract from our primary findings.

## Conclusions

Incomplete imaging surveillance following EVAR remains a significant problem and predisposes patients to worse survival outcomes. Within a statewide quality collaborative, hospital-specific reporting of EVAR surveillance rates and an associated pay-for-performance metric, is a viable strategy that can have a significant impact on post-EVAR 1-year surveillance imaging and decreasing long-term mortality after endovascular repair.

## Data Availability

The data for this manuscript can be available upon request to the corresponding author

---

**Supplemental Figure 1. All risk factors analyzed for repeat procedure**.

**Supplemental Figure 2. All risk factors analyzed for patient mortality**.

